# Mental Bandwidth is Associated with HIV and Viral Suppression Among Low-Income Women in Philadelphia

**DOI:** 10.1101/2024.03.25.24304870

**Authors:** Aaron Richterman, Nancy Aitcheson, Celeste Durnwald, Cara Curley, William R. Short, Mirabelle Jean Louis, Florence Momplaisir, Harsha Thirumurthy

**Author notes:** Corresponding Author:* Aaron Richterman, MD, MPH; Hospital of the University of Pennsylvania, 3400 Spruce Street, Philadelphia, PA 19104.

## Abstract

Behavioral economics research suggests poverty may influence behavior by reducing mental bandwidth, increasing future discounting, and increasing risk aversion. It is plausible that these decision-making processes are further impaired in the context of HIV or pregnancy. In this cross-sectional study of 86 low-income women in Philadelphia, multivariable models showed that HIV was associated with decreased mental bandwidth (one of two measures) and lower risk aversion. Pregnancy was not associated with any decision-making factors. Viral suppression was associated with greater mental bandwidth (one of two measures), and antenatal care engagement with lower future discounting. Anti-poverty interventions may be particularly beneficial to improve health behaviors in the context of HIV.

## Introduction

Health behaviors such as lack of adherence to antiretroviral therapy (ART) and inconsistent engagement in care remain some of the greatest barriers to ending HIV epidemics. This is particularly true for pregnant women with HIV, for whom postpartum clinic attendance, ART adherence, and postpartum viral suppression rates remain unacceptably low in high– and low-income settings.^1-10^

40% of people with HIV (PWH) in the US live below the poverty line,^11^ and poverty is an important upstream contributor to poor ART adherence and engagement in care by PWH, particularly during pregnancy and in the years after giving birth.^5,9^ Recent behavioral economics research suggests that poverty may affect individual behavior through psychological (in addition to economic) mechanisms.^12-21^ This may occur through decreases in mental bandwidth (the mental resources available at a given time to make complex decisions), increases in future discounting (the tendency to undervalue future outcomes), and increases in risk aversion.^15,16,19-24^

While the association between poverty and these psychological decision-making processes has been recently established, it is plausible that these processes are even further impaired in the context of HIV or pregnancy. If this were the case, it would suggest that anti-poverty interventions could be particularly beneficial to improve health behaviors in these populations. To evaluate this possibility, we measured mental bandwidth, future discounting, and risk aversion among low-income women in Philadelphia, and examined their associations with HIV and pregnancy status.

## Methods

We conducted a cross-sectional study of low-income pregnant and non-pregnant women with and without HIV receiving care at outpatient clinics in the University of Pennsylvania Health System. Eligibility criteria included (1) cisgender female, (2) age 18-40 years, (3) Philadelphia resident, (4) household income <200% federal poverty level, (5) confirmed pregnancy by standard laboratory and ultrasound testing (pregnant participants), (6) documented HIV seropositive (participants with HIV), (7) able to provide informed consent. The study was approved by the University of Pennsylvania IRB. All participants provided written informed consent.

Participants completed a one-time survey assessing sociodemographics, income, food security,^25^ health insurance, obstetrical history, self-reported ART adherence (among PWH), and whether a pregnancy was intended (pregnant participants). We conducted chart reviews to assess for most recent HIV viral load (last 3 months) and current gestational age. For pregnant participants, we conducted a follow-up chart review after delivery to assess antenatal care engagement (proportion of scheduled obstetrics clinic appointments that were attended) and gestational age at delivery.

We assessed mental bandwidth using two measures.^14^ The Psychomotor Vigilance Task (PVT) assesses attentional vigilance by asking participants to press a button when a stimulus appears on a screen over a period of ten minutes and measuring reaction time and accuracy.^26,27^ After standardizing reaction time, number of false starts (responding prior to the stimulus appearing), and number of minor lapses (correctly responding but with response time >500 milliseconds) such that each variable had a population mean of 0 and standard deviation (SD) of 1, we generated PVT scores to synthesize reaction time and accuracy as follows: reaction time + 0.5*(minor lapses + false starts).^28^ Raven’s Progressive Matrices (RPM) is a non-verbal estimate of fluid intelligence, or the ability to solve novel problems or adapt to new situations.^29^ RPM presents participants with a series of ten figures with a missing portion of a pattern and asks them to select which of six options best completes the missing pattern, with the number of correct answers representing a participant’s score.

We assessed future discounting using the Kirby Delay-Discounting Task (KDDT), a 27-item questionnaire that asks participants to hypothetically choose between smaller, immediate monetary rewards and larger, delayed awards.^30,31^ The KDDT generates each participant’s discounting constant (*k*), with higher values indicating greater amounts of future discounting (i.e., greater impulsivity).^32^

We assessed risk-aversion using an ordered lottery selection method, asking participants hypothetical questions about whether they would prefer a 100% chance of receiving $5, or a 50% chance of receiving $10 and a 50% chance of receiving $0, $1, $2, $3, or $4, to assess the threshold (if any) at which they would choose the higher-risk option.^33,34^

We used multivariable linear regression models to separately estimate associations between HIV and pregnancy (primary explanatory variables) with mental bandwidth (PVT, RPM), future discounting, and risk aversion (primary outcomes). Each of the outcomes was standardized to have a mean of 0 and standard deviation (SD) of 1, such that effect sizes were expressed in SDs of the outcome. Models controlled for age, race, ethnicity, educational attainment, income, food security, pregnancy (HIV models), and HIV (pregnancy models). These covariates were selected a priori as either known or hypothesized confounders of the relationship between the decision-making processes and HIV or pregnancy.

In secondary analyses, we used similar multivariable models to estimate associations between the same decision-making factors and viral suppression (among PWH) and antenatal care engagement (among pregnant participants). Because of smaller sample sizes, these models adjusted only for age and either HIV or pregnancy.

We had a planned sample size of at least 70 participants (approximately half with HIV, and half pregnant) to provide at least 80% power to detect similar differences in mental bandwidth as seen in prior research demonstrating the effects of poverty alleviation on mental bandwidth (0.67 SDs).^15^ The study was approved by the University of Pennsylvania Institutional Review Board. We performed statistical analysis using SAS Version 9.4 (SAS Institute; Cary, NC).

## Results

We enrolled 86 participants with a median age of 28 years (IQR 25 to 32) between March 2022 and August 2023. There were 69 (80%) participants who identified as non-Hispanic Black, 6 (7%) as non-Hispanic White, 2 (3%) as non-Hispanic Asian, 2 (3%) as non-Hispanic and multiple races, and 7 (8%) as Hispanic. Sixty-seven participants (78%) had a single marital status, and 76 (88%) had completed high school. Participants had a median of 1 child (IQR 1 to 2). For annual household income, 25 participants (29%) reported ≤$10,000, 20 (23%) reported $10,001-$20,000, 28 (33%) reported $20,001-$40,000, 10 (12%) reported $40,001-$60,000, and 3 (3%) reported $60,001-$80,000. Fifty-four participants (63%) were employed, 33 (38%) had low or very low food security, and 76 (88%) had Medicaid.

Thirty-five participants (41%) were living with HIV, and 36 (42%) were pregnant — 26 (30%) were non-pregnant without HIV, 24 (28%) were non-pregnant with HIV, 25 (29%) were pregnant without HIV, and 11 (13%) were pregnant with HIV. PWH reported a median of one (IQR 0 to 3) missed dose of ART during the last month, with 16 (46%) having <95% adherence and 4 (11%) having <80% adherence. The most recent viral load was undetectable for 30 (86%) of the PWH. Among pregnant participants, 9 (25%) intended the current pregnancy, and the median gestational age at the time of the enrollment was 25 weeks (IQR 16 to 32). In follow-up chart review, pregnant participants had a median engagement in antenatal care (proportion of scheduled obstetrics visits attended) of 0.86 (IQR 0.71 to 1.00), and a median gestational age at delivery of 39 weeks (IQR 37 to 40).

Participants had a median number of correct answers on the RPM of 8 (IQR 7 to 9), and on the PVT they had a median reaction time of 414 milliseconds (IQR 375 to 484), median number of false starts of 1 (IQR 0 to 2), and a median number of minor lapses of 17 (IQR 8 to 38). Participants had a median log temporal discounting constant of –3.7 (IQR –4.6 to –2.7). The median risk-aversion threshold was a 50% chance of $4 and a 50% chance of $10 (IQR 50% chance of $1 to no threshold [always choosing a 100% chance of $5]).

After adjustment, HIV was associated with lower mental bandwidth on the PVT (−0.42 SD difference, 95% CI –0.84 to –0.01), and lower risk-aversion (−1.16 SD difference, 95% CI –2.05 to –0.27) (Table 2). Pregnancy was not associated with any decision-making factors. Among PWH, viral suppression was associated with greater mental bandwidth on the PVT (1.12 SD difference, 95% CI 0.10 to 2.13). Among pregnant participants, antenatal care engagement was associated with lower future discounting (−1.57 SD difference, 95% CI –3.08 to –0.05).

## Discussion

In this cross-sectional study of low-income women receiving health care in Philadelphia, we found that living with HIV was associated with significantly lower mental bandwidth (on one of two measures) and risk aversion, while pregnancy was not associated with any psychological decision-making factors. We also found that viral suppression was associated with greater mental bandwidth (on one of two measures), and antenatal care engagement was associated with lower future discounting. The differences were largely similar in magnitude to those seen in prior studies of poverty alleviation.^15^ We hypothesize that we found associations between HIV (and viral suppression) and mental bandwidth when using the PVT but not RPM because of a ceiling effect resulting from high scores with little variability for the RPM, indicating that the RPM series needs to be more difficult for this population in future work. This is among the first studies to assess these decision-making factors among low-income PWH and pregnant women.

Our findings suggest that PWH living in poverty may be especially constrained in their ability to make decisions to promote long-term health (e.g. medication adherence), and that these constraints are associated with important health outcomes like viral suppression and engagement in care. Consequently, interventions that increase mental bandwidth and decrease future discounting, such as poverty alleviation through cash transfers or housing support, may lead to improvements in health behaviors and health outcomes. While we did not find any specific differences associated with pregnancy, it remains possible that sub-groups of pregnant women – such as those with limited social support – experience greater losses in mental bandwidth. Studies with larger sample sizes of low-income pregnant women could further explore such associations. Our findings nonetheless remain relevant during the perinatal period since poverty often increases around the time of pregnancy.^35^

This study expands our understanding of poverty’s impact on psychological decision-making processes into the context of HIV and HIV-related behaviors. The psychology literature points to a dual-process model of thinking and decision-making — System 1 is intuitive, and rapidly guides one’s decision-making on a regular basis; System 2 is deliberative and considers longer-term consequences of each decision, but requires cognitive effort.^36^ Mental bandwidth has been defined as the mental resources available at a given time to engage in System 2 thinking.^14^ Recent behavioral economics research has shown that a state of economic scarcity (i.e., living in poverty) induces people to focus most of their attention towards immediate needs,^19,23,24^ and that attention to these needs diminishes bandwidth.^15,20^ Similarly, evidence suggests that poverty affects time preferences by increasing future discounting (defined as the tendency to undervalue future outcomes).^21^ In contrast, alleviating scarcity through poverty alleviation has been shown to increase bandwidth and decrease discounting.^16,22^ Whether poverty alleviation’s effects on mental bandwidth and future discounting translates into changes in future-oriented health behaviors remains an open question. While this growing economics literature has focused on the effects of poverty on these psychological decision-making factors, our study is among the first to evaluate associations between these factors and a specific context like HIV within which poverty is a highly important determinant of outcomes.

This study has several limitations. First, this study was cross-sectional, which precludes establishing causality for the identified associations. For example, living with HIV may lead to reductions in mental bandwidth, but it’s also possible that people with lower mental bandwidth (because of poverty and other factors) may be more likely to acquire HIV, or that lower mental bandwidth is co-correlated with other HIV risk factors (such as substance use or housing insecurity). Regardless, the fact that PWH have lower mental bandwidth indicates that they may especially benefit from economic interventions that increase bandwidth and potentially improve health behaviors like ART adherence.^37^ Second, as our findings come from a single center in a high-income country, they should be considered preliminary and replicated in other contexts.

## Conclusion

In this cross-sectional study of low-income women living in Philadelphia, living with HIV was associated with lower mental bandwidth (on one of two measures) and risk aversion, whereas pregnancy was not associated with any psychological decision-making factors. Our findings suggest that PWH living in poverty may be especially constrained in their ability to make daily decisions to promote long-term health. We also found that these constraints are associated with important health outcomes like viral suppression and engagement in care. Consequently, interventions that increase mental bandwidth and decrease future discounting, such as poverty alleviation, may lead to important improvements in outcomes among PWH.

## Data Availability

All data produced in the present study are available upon reasonable request to the authors

## Abbreviations

ART: antiretroviral therapy
HIV: human immunodeficiency virus
IQR: interquartile range
KDDT: Kirby Delay Discounting Task
PWH: people with HIV
PVT: psychomotor vigilance task
RPM: Raven’s progressive matrices
SD: standard deviation

## Author Roles

### Author contributions

Richterman had full access to all data in the study and takes responsibility for the integrity of the data and accuracy of the data analysis.

### Concept and design

Richterman, Aitcheson, Momplaisir, Thirumurthy

### Acquisition, analysis, interpretation of data

All authors

### Drafting of the manuscript

Richterman

### Critical revision of the manuscript

All authors

### Statistical analysis

Richterman

---

**Table 1.** Associations (with 95% CIs) between mental bandwidth, risk aversion, and future discounting with HIV, viral suppression (among PWH), pregnancy, and antenatal care engagement (among pregnant participants). Effect measures are generated using multivariable linear regression models and are and standardized such that effect sizes are expressed in standard deviations of the outcomes. HIV and pregnancy models control for age, race, ethnicity, educational attainment, income, food security, pregnancy (HIV model), and HIV (pregnancy model). The viral suppression and antenatal care engagement models control for age and HIV or pregnancy. Sample sizes are as follows: HIV and Pregnancy models N=86; viral suppression model N=35; antenatal care model N=36.

|  | HIV<sup>1</sup><br>(N=86) | Viral Suppression<br>(among PWH)<sup>2</sup><br>(N=35) | Pregnancy<sup>3</sup><br>(N=86) | Antenatal care<br>engagement (among<br>pregnant)<sup>4</sup><br>(N=36) |
| --- | --- | --- | --- | --- |
| Mental<br>Bandwidth<br>(Psychomotor<br>Vigilance<br>Task) | -0.42 (-0.84 to -0.01) | 1.12 (0.10 to 2.13) | 0.09 (-0.31 to 0.49) | 0.89 (-0.68 to 2.46) |
| Mental<br>Bandwidth<br>(Raven's<br>Progressive<br>Matrices) | -0.37 (-0.78 to 0.06) | -0.48 (-1.59 to 0.63) | 0.07 (-0.34 to 0.47) | -0.32 (-1.79 to 1.15) |
| Future<br>Discounting | -0.07 (-0.51 to 0.37) | -0.11 (-1.10 to 0.89) | -0.03 (-0.45 to<br>0.39) | -1.57 (-3.08 to -0.05) |
| Risk aversion | -1.16 (-2.05 to -0.27) | 0.08 (-1.72 to 1.88) | -0.62 (-1.48 to<br>0.23) | -0.16 (-3.20 to 2.88) |

<sup>1</sup> Models control for age, race, ethnicity, educational attainment, income, food security, and pregnancy.
<sup>2</sup> Models control for age and pregnancy.
<sup>3</sup> Models control for age, race, ethnicity, educational attainment, income, food security, and HIV status.
<sup>4</sup> Models control for age and HIV or pregnancy.

